# Public Reactions towards the COVID-19 Pandemic on Twitter in the United Kingdom and the United States

**DOI:** 10.1101/2020.07.25.20162024

**Authors:** Canruo Zou, Xueting Wang, Zidian Xie, Dongmei Li

## Abstract

**Background:** The coronavirus disease 2019 (COVID-19) has spread globally since December 2019. Twitter is a popular social media platform with active discussions about the COVID-19 pandemic. The public reactions on Twitter about the COVID-19 pandemic in different countries have not been studied. This study aims to compare the public reactions towards the COVID-19 pandemic between the United Kingdom and the United States from March 6, 2020 to April 2, 2020.

**Data:** The numbers of confirmed COVID-19 cases in the United Kingdom and the United States were obtained from the 1Point3Acres website. Twitter data were collected using COVID-19 related keywords from March 6, 2020 to April 2, 2020.

**Methods:** Temporal analyses were performed on COVID-19 related Twitter posts (tweets) during the study period to show daily trends and hourly trends. The sentiment scores of the tweets on COVID-19 were analyzed and associated with the policy announcements and the number of confirmed COVID-19 cases. Topic modeling was conducted to identify related topics discussed with COVID-19 in the United Kingdom and the United States.

**Results:** The number of daily new confirmed COVID-19 cases in the United Kingdom was significantly lower than that in the United States during our study period. There were 3,556,442 COVID-19 tweets in the United Kingdom and 16,280,065 tweets in the United States during the study period. The number of COVID-19 tweets per 10,000 Twitter users in the United Kingdom was lower than that in the United States. The sentiment scores of COVID-19 tweets in the United Kingdom were less negative than those in the United States. The topics discussed in COVID-19 tweets in the United Kingdom were mostly about the gratitude to government and health workers, while the topics in the United States were mostly about the global COVID-19 pandemic situation.

**Conclusion:** Our study showed correlations between the public reactions towards the COVID-19 pandemic on Twitter and the confirmed COVID-19 cases as well as the policies related to the COVID-19 pandemic in the United Kingdom and the United States.

## Introduction

A novel coronavirus disease, known as COVID-19, was identified in China in December 2019 [1]. This virus can transmit from person to person, and the most common symptoms include fever, dry cough, and tiredness [2]. On March 11, 2020, the World Health Organization declared the outbreak a pandemic [3]. As of June 16, 2020, there have been around 8 million COVID-19 cases worldwide [4]. The United States had the first confirmed cases on January 22, 2020, and the United Kingdom had the first confirmed cases on January 31, 2020 [4]. These two countries had their first COVID-19 case in late January, but by June 14, the United States (2,141,057 COVID-19 cases) had around 7 times confirmed cases of that in the United Kingdom (298,139 cases) [4]. Thus, it is important to examine the differences in public reactions to this COVID-19 pandemic between these two countries.

Since the outbreak of H1N1 (a novel influenza A virus) in 2009, the internet has been the most frequently used source of information for the public to learn about the pandemic [5]. Social media platforms provide unique resources where people can access and share information. Twitter is a social media platform with 330 million monthly active users, where the users can post brief (<280 characters) text messages known as “tweets” [6]. Mining the data on Twitter can help us understand the public’s opinions and behavioral responses to the COVID-19 pandemic.

Most of social media studies on COVID-19 focused on one particular country [7], misinformation [8]–[10], or mental health [11]. A cross-region comparison study analyzed the Twitter contents about COVID-19 of world leaders in G7 countries, but they did not evaluate its association with the public [12]. To our best knowledge, attempts to discover public reactions to COVID-19 on social media across countries are still lacking. In this study, statistical analysis and text mining techniques were applied to the COVID-19 related tweets to understand how Twitter users responded to the COVID-19 pandemic in the United Kingdom and the United States. Our findings suggest that Twitter users in different countries have different reactions towards the COVID-19 pandemic, which may be related to the severity of COVID-19 pandemic and related policies/news in different countries.

## Methods

### Data collection

The number of confirmed COVID-19 cases in the United Kingdom and the United States from March 6, 2020 to April 2, 2020 were obtained from the 1Point3Acres website [4]. The Twitter dataset used for this study was generated by a crawler using the Twitter streaming API from March 6, 2020 to April 2, 2020 [13]. A set of keywords, “CORONA”, “corona”, “COVID19”, “covid19”, “covid”, “coronavirus”,”Coronavirus”, “CoronaVirus”, and “NCOV”, was used to collect COVID-19 related tweets. The total number of COVID-19 tweets in the dataset was 155,028,779.

### Data preprocessing

Using a Python script, the tweets without COVID-19 related keywords in the text were filtered out. There is a beer named Corona Extra produced by Cervecería Modelo. The tweets mentioned corona as beer had been filtered out. Another set of keywords, “dealer”, “deal”, “supply”, “beer”, “drink”, “drank”, “drunk”, “store”, “promo”, “promotion”, “customer”, “discount”, “sale”, “free shipping”, “sell”, “$”, “%”, “dollar”, “offer”, “percent off”, “save”, “price”, “wholesale”, was used to filter out unrelated and promotion or commercial tweets. After filtering out tweets irrelevant to COVID-19, the dataset contained 85,953,249 tweets.

The names of the country, state, and cities with the top 50 population, such as “New York, New York” for New York City, and “California” for California state, were used to filter tweets in the United States. The names of the country such as England, Scotland, Wales, and Northern Ireland, and cities with the top 20 population were used to filter tweets in the United Kingdom. After filtering the data by the geolocation of Twitter users, the number of COVID-19 tweets from the United States was 19,942,119 while the number of COVID-19 tweets from the United Kingdom was 4,459,655.

A Python script was used to check if some tweets with the same text were posted by the same user so that the duplicate tweets were removed. The number of unique COVID-19 tweets in the United States was 16,280,065 and the number of unique tweets in the United Kingdom was 3,556,442, which were used for the final analysis.

Because the raw tweets contained the tweet created time in the Coordinated Universal Time (UTC), the tweet created time was converted to the respective local time zones for each region to facilitate the comparison. The data in the United States were converted to UTC-08:00. The data in the United Kingdom were converted to UTC+01:00.

### Temporal analysis

A temporal analysis was performed to investigate the daily and hourly change in the number of COVID-19 tweets during the study period. The temporal trend of the number of tweets per 10,000 Twitter users during the study period was calculated by the daily number of tweets normalized by the number of users in the region. We also performed the temporal hourly trend of the COVID-19 tweets by calculating the average number of tweets by hour, and then normalized by the total number of users in the region.

### Sentiment analysis

Sentiment analysis was conducted to examine the public attitudes toward the COVID-19 pandemic in both the United Kingdom and the United States. The sentiment analysis tool Valence Aware Dictionary and sEntiment Reasoner (VADER) was used to calculate the sentiment score for each tweet [14]. The word “virus” was removed from the text before sentiment analysis because this study focused on public attitudes about the virus and this word itself could influence the sentiment.

To analyze the sentiment during the study period, VADER was applied to the dataset, which calculated the daily average sentiment scores of COVID-19 tweets in the United Kingdom and the United States. To analyze the sentiment scores in a 24-hour period, VADER calculated the hourly average sentiment scores of all COVID-19 tweets.

### Topic modeling

A tool for Latent Dirichlet Allocation (LDA) topic models, Gensim parallelized LDA, was applied to identify the COVID-19 related topics on Twitter in the United Kingdom and the United States [15]. Due to the time-consuming training process of LDA models, 30% of tweets were randomly sampled from the datasets. Before the model was built, the text was split into sentences and further split into words. All characters were converted to the lowercase to ensure consistency. Unrelated characters, including emails, newline, extra spaces, distracting single quotes, and URLs, were removed from the data. Stopwords from the Natural Language ToolKit and the words “virus”, “corona”, “coronavirus”, “ncov”, “covid19” and “covid” were also removed due to unrelatedness [16]. Words were lemmatized and stemmed to their root forms using spaCy because different forms of the same words have the same meaning in this study [17].

LDA is unsupervised, and prior to running the model, the number of topics exits in the corpus is unknown. Models of different numbers of topics were built and the model with the best performance was chosen to represent the data. Intertopic distance map using pyLDAVis and topic coherence using Genism were used to evaluate the model. In the intertopic distance map, topics were plotted as circles in the two-dimensional plane whose centers are determined by the computed distance between topics [18]. If two circles overlapped each other, the corpus could have been modeled with fewer topics. The topic coherence score measures a single topic by measuring the degree of semantic similarity between high scoring words in the topic. A higher topic coherence score indicates a better model. Through an iterative process, the United Kingdom data had the highest coherence score with 10 topics, and the United States data had the highest coherence score with 12 topics.

## Results

### Temporal analysis of COVID-19 cases and related tweets in the United Kingdom and the United States

Figure 1 showed the daily confirmed new COVID-19 cases in the United Kingdom and the United States from March 6 to April 2, 2020. The numbers of COVID-19 cases were low in both countries before March 18, 2020 with similar magnitude. After March 18, 2020, the rate of increase in new COVID-19 cases in the United States was significantly higher than that in the United Kingdom while the number of new COVID-19 cases in both countries increased. On April 2, 2020, the newly confirmed COVID-19 cases in the United States (30,081 cases) were around seven times of that in the United Kingdom (4,244 cases).

**Figure 1.**
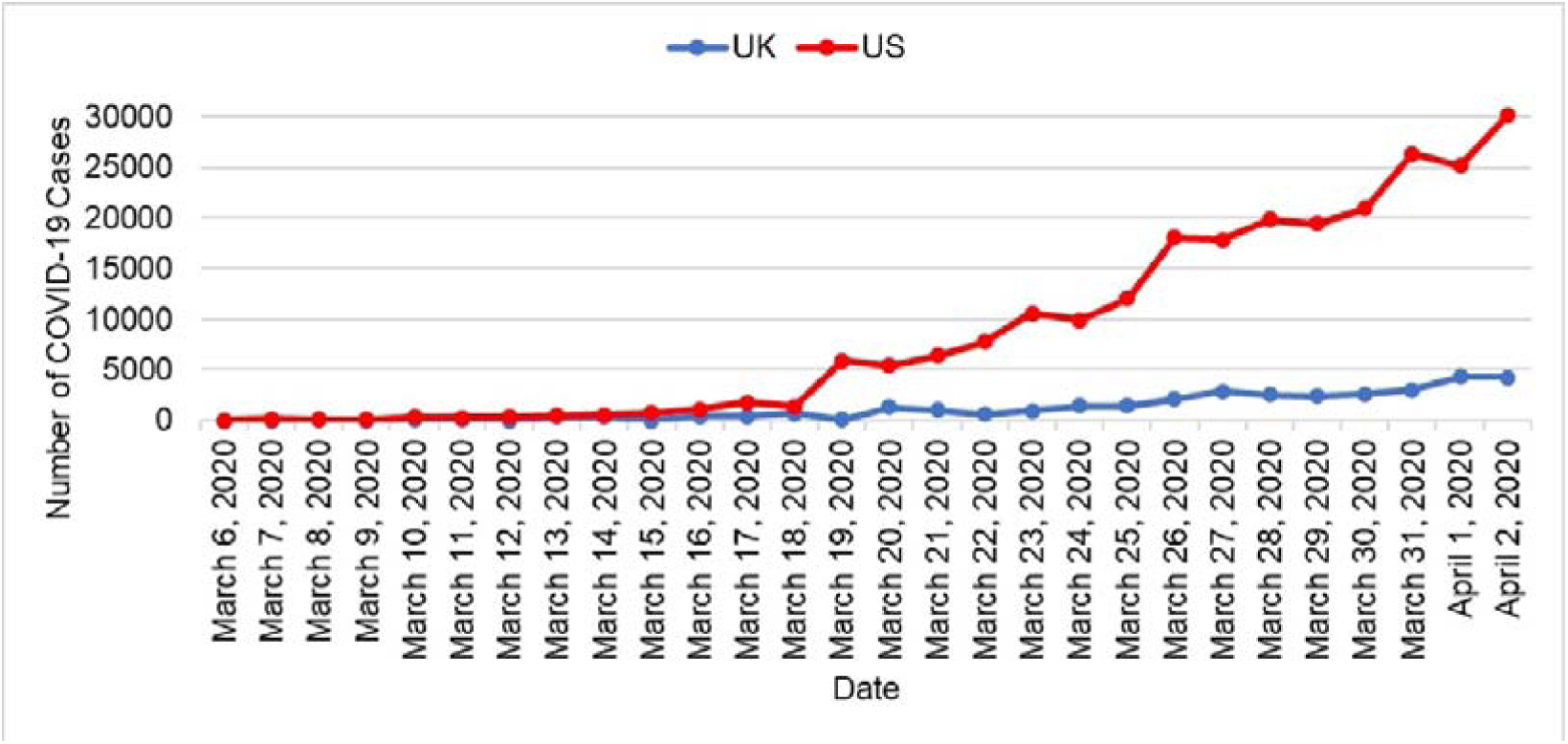
Daily confirmed new COVID-19 cases in the United Kingdom (UK) and the United States (US).

The temporal trend of the COVID-19 related tweets between the two countries was different (Figure 2). Overall, the number of COVID-19 tweets per 10,000 twitter users in the United States was higher than that in the United Kingdom. The numbers of COVID-19 tweets in the United Kingdom fluctuated between March 6 and April 2, 2020, while the numbers of tweets in the United States decreased significantly after March 11.

**Figure 2.**
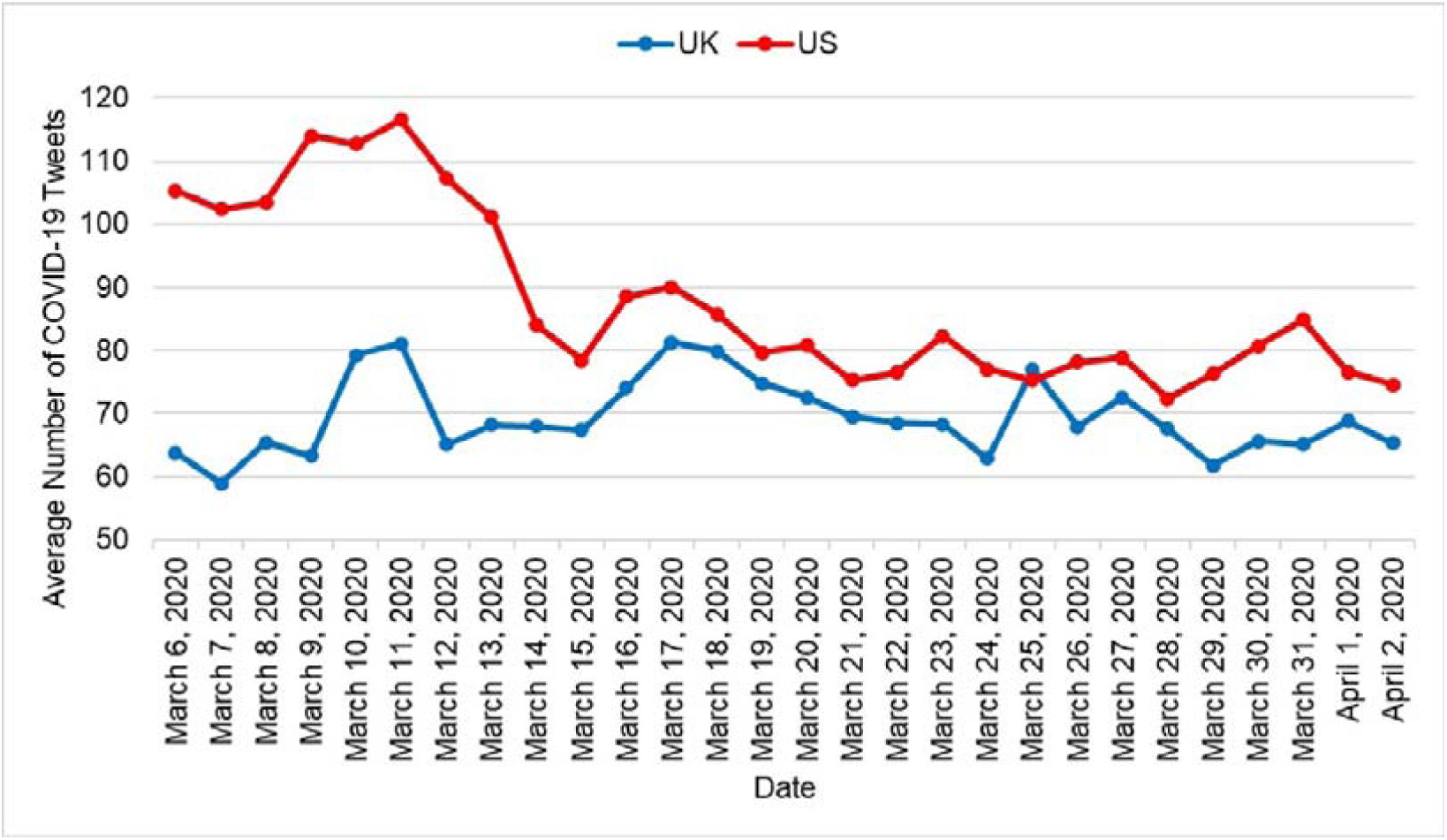
The number of COVID-19 related tweets per 10,000 Twitter users over time in the United Kingdom (UK) and the United States (US).

### Public attitudes towards the COVID-19 pandemic in the United Kingdom and the United States

The sentiment scores of COVID-19 related tweets between the United Kingdom and the United States had different patterns before March 18 but similar patterns after March 18, 2020 (Figure 3). There was a large drop in the sentiment score in both countries from March 20 to March 23, 2020. The sentiment scores in the United States were lower than those in the United Kingdom except on March 8. The sentiment score on March 25, 2020 in the United Kingdom was positive (≥ 0.05), while all others were neutral or negative.

**Figure 3.**
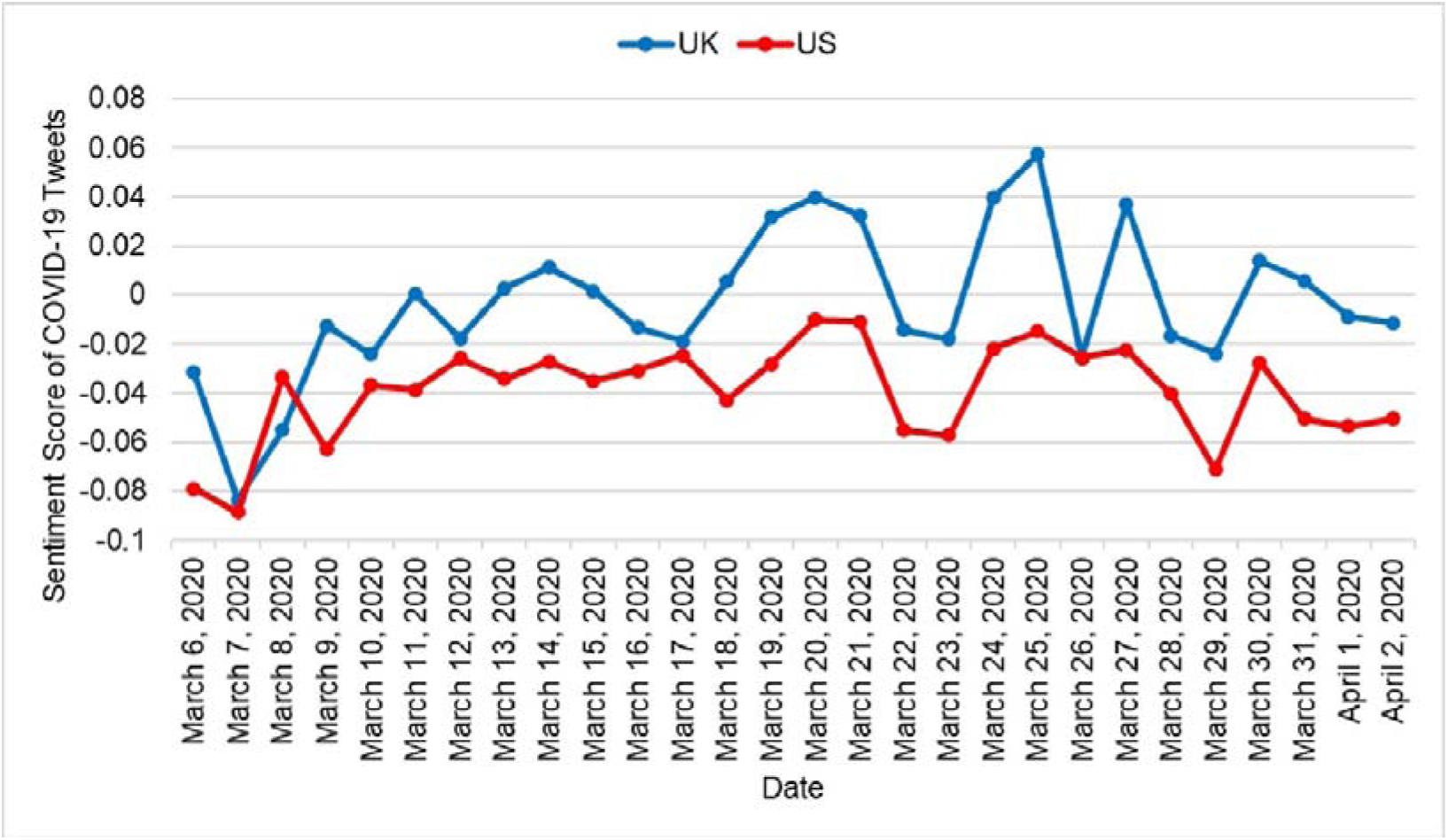
Sentiment scores of COVID-19 related tweets over time in the United Kingdom (UK) and the United States (US).

The number of COVID-19 tweets in the United Kingdom and the United States had different patterns in a 24-hour period (Figure 4A). In the United Kingdom, the number of COVID-19 tweets increased from 5 AM until 9 AM, and then slowly dropped. In contrast, in the United States, the number of COVID-19 tweets started to increase slowly from 3 AM and reached a peak between 4 PM and 6 PM.

**Figure 4.**
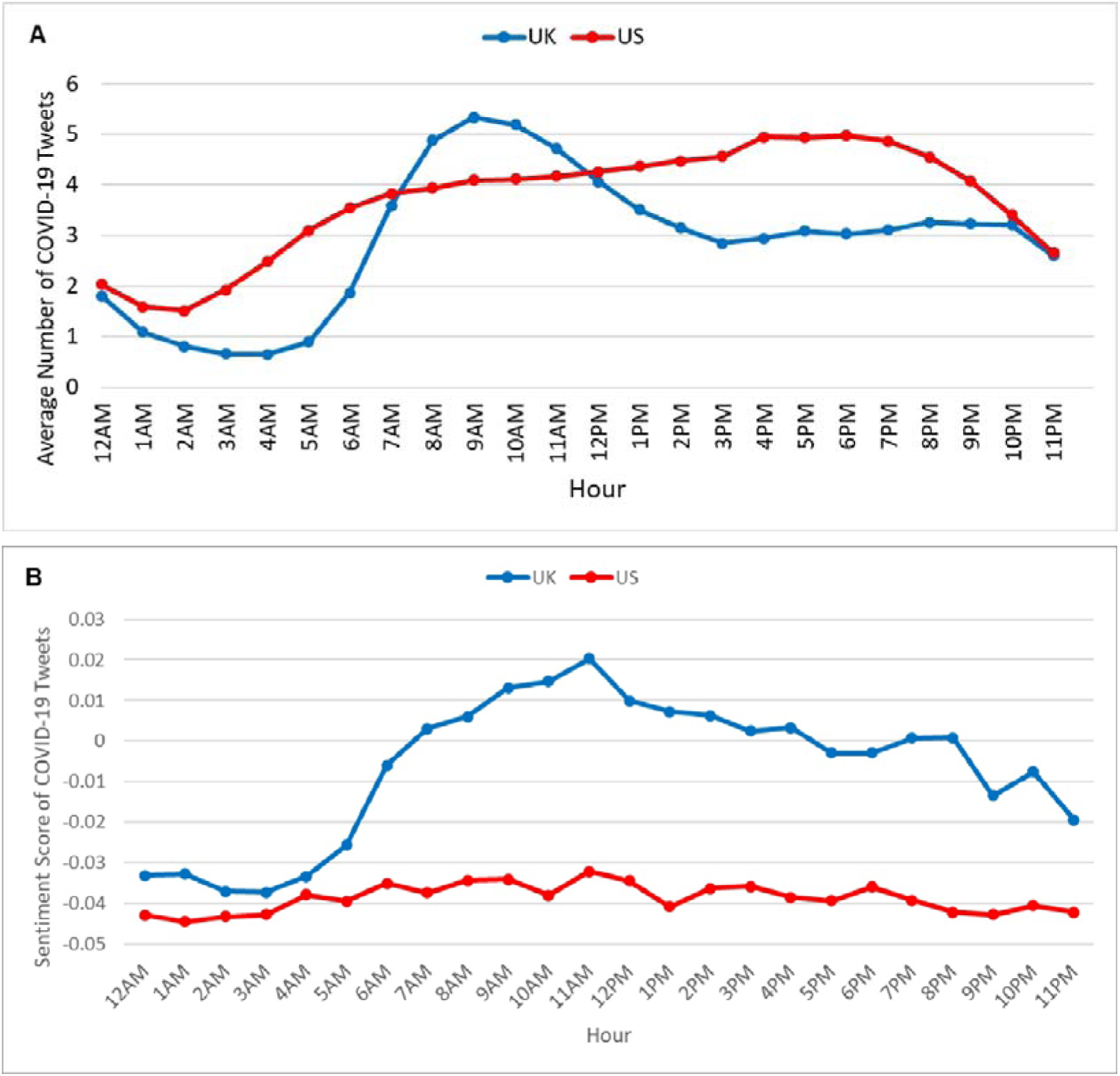
The number of COVID-19 tweets and sentiment scores in a 24-hour period in the United Kingdom (UK) and the United States (US). (A) The number of COVID-19 tweets; (B) Sentiment scores of COVID-19 tweets.

The sentiment scores of COVID-19 related tweets in the United Kingdom and the United States had different patterns in a 24-hour period (Figure 4B). The sentiment scores towards the COVID-19 pandemic in the United States were always lower than those in the United Kingdom. The sentiment scores remained relatively constant over time in the United States. In contrast, the sentiment scores in the United Kingdom started to increase from 3 AM and reached a peak at 11 AM, and then slowly decreased.

### Top topics associated with COVID-19 in the United Kingdom and the United States

Based on LDA modeling, there were 10 topics discussed on Twitter related to COVID-19 in the United Kingdom and 12 topics discussed in the United States (Table 1). Both the United Kingdom and the United States had topics about COVID-19 cases and deaths, and topics about social distancing. The COVID-19 tweets in the United Kingdom had unique topics about gratitude and support, while the tweets in the United States had unique topics about the pandemic and government.

**Table 1.** Top topics of COVID-19 related Tweets in the United Kingdom (UK) and the United States (US)

| US |  |  | UK |  |  |
| --- | --- | --- | --- | --- | --- |
| Topics | % Tweets | Top 10 Keywords | Topics | % Tweets | Top 10 Keywords |
| global pandemic situation | 10.81% | pandemic, time, end, lose, great, fight, global, Trump, use, world | gratitude to government and health workers | 14.87% | health, thank, government, worker, care, public, late, support, read, help |
| government and positive cases | 10.42% | test, positive, government, say, official, member, doctor, today, social, patient | new cases and deaths | 11.79% | test, case, new, death, positive, number, report, day, lockdown, today |
| people's death | 9.32% | die, say, people, get, year, start, kill, day, many, flu | family support | 10.12% | help, support, know, even, family, need, question, right, share, child |
| need support from the medical system | 9.00% | need, order, hospital, worker, patient, health, care, leave, testing, pay | food security | 9.63% | make, time, give, people, look, think, want, food, get, run |
| new cases and deaths | 8.94% | case, death, new, report, first, state, confirm, number, day, today | death in patients | 8.43% | die, hospital, patient, last, year, symptom, hour, old, day, take |
| school closure | 8.90% | get, go, life, think, close, make, people, school, country, really | consequence of COVID-19 | 8.29% | get, people, say, many, can, lose, die, young, bad, contract |
| health care and political event | 8.27% | call, know, take, s, come, go, doctor, impeachment, seriously, wait | work from home | 7.94% | work, home, amp, go, be, stop, stay, doctor, staff, nurse |
| federal government | 8.01% | Trump, relief, month, let, question, medium, check, vote, free, get | self-isolation | 6.89% | week, pandemic, self, day, come, live, back, friend, isolation, isolate |
| conference cancellation | 7.43% | due, right, cancel, week, hold, conference, press, hour, panic, Chinese | hope remains | 6.47% | due, crisis, really, still, close, let, hope, kill, cancel, happen |
| Trump's statement on drugs | 5.53% | make, crisis, Trump, away, drug, lie, death, minute, say, find | fight COVID-19 to protect jobs | 5.91% | fight, use, spread, help, put, risk, protect, part, government, job |
| work from home | 3.84% | home, work, news, stay, feel, put, people, say, go, never |  |  |  |
| protective measures | 3.56% | amp, stop, be, mask, hand, go, people, early, work, wash |  |  |  |

## Discussion

To understand how the severity of COVID-19 pandemic and related policies affect the public reactions towards the COVID-19 pandemic on social media, we collected and analyzed the COVID-19 related tweets from March 6 to April 2, 2020 coming from the United Kingdom and the United States. Our results showed that the number of confirmed COVID-19 cases in the United Kingdom was significantly less than that in the United States. Similarly, the number of COVID-19 related tweets posted by every 10,000 Twitter users in the United Kingdom was lower than that in the United States, which indicates that COVID-19 was a less popular topic on Twitter in the United Kingdom than that in the United States. In addition, the daily sentiment scores of COVID-19 related tweets in the United Kingdom were higher than those in the United States in general, which suggests that the COVID-19 related tweets in the United Kingdom had a more positive attitude towards COVID-19 than those in the United States. Moreover, the United Kingdom had a large proportion of topics with positive attitudes, such as gratitude and support, while in the United States, a large proportion of topics were about the pandemic and cases. This might be due to the smaller number of daily new COVID-19 cases in the United Kingdom than in the United States.

While the sentiments scores towards the COVID-19 pandemic in the United States were more negative than those in the United Kingdom, the daily trends of the number of COVID-19 tweets and their corresponding sentiment scores in these two countries fluctuated. This result may be that the Twitter users were influenced by the announcement of major policies related to COVID-19 and some celebrities tested positive for COVID-19. For example, while the World Health Organization declared the outbreak a pandemic on March 11, 2020 [3], and the United Kingdom government announced £330 billion of loans to support businesses on March 17, 2020 [19], the number of COVID-19 tweets had peaks in both the United Kingdom and the United States. The sentiment scores towards COVID-19 in both the United Kingdom and the United States had peaks around March 21, 2020, when New York, California and other large states in the United States had ordered most businesses to close by March 21, 2020 [20], and British PM Johnson requested entertainment venues to close on March 20, 2020 [21]. The number of COVID-19 tweets was high in the United Kingdom when Prince Charles tested positive for the COVID-19 on March 25, 2020 [22], and Prime Minister Boris Johnson tested positive for the COVID-19 on March 27, 2020 [23]. Likewise, the number of COVID-19 tweets was high on March 23 and March 31, 2020 in the United States when the number of new confirmed COVID-19 cases in the United States was high in these two days [4]. Therefore, the news about celebrities tested positive with COVID-19 and a historically high number of new COVID-19 confirmed cases can motivate Twitter users’ discussion. The sentiment scores of COVID-19 tweets in the United Kingdom had two peaks on March 25 and March 27, 2020, and then dropped dramatically one day after. The reason may be that the Twitter users had some lay back on the negative emotion about the celebrities tested positive for COVID-19. The sentiment scores in both countries dropped on March 22, 2020, which might result from that the Twitter users in both countries were unhappy after the entertainment facilities were closed. Nguyen et al. proposed a stochastic burst detection model to extract sentimental bursts from blogs, and the topics extracted from the bursts matched with real-world news events [24]. In this study, we demonstrated that the number of COVID-19 related tweets and their sentiment scores were correlated with the significant events about COVID-19.

In this study, we showed that the sentiment scores of COVID-19 related tweets had a diurnal pattern. Dzogang et al. showed that two independent factors could explain 85% of the variance across tweets’ 24-hour profiles in the United Kingdom, one starting at 5 AM/6 AM correlates with positive emotions and another starting at 3 AM/4 AM correlates with negative affect and social concerns [25]. Consistently, our study showed that the sentiment scores of COVID-19 related tweets in the United Kingdom dropped to the bottom at 3 AM and started to increase rapidly from 5 AM. In addition, we showed that the number of COVID-19 tweets in the United Kingdom had a diurnal pattern, in which the number of COVID-19 tweets was the highest at 9 AM and the lowest at 3 AM. The reasons behind this observation need further investigation. As for the United States, Golder et al. showed that there was a morning rise and night-time peak in positive affect, and a sharp drop in negative affect during the overnight hours on Twitter [26]. We found a similar pattern in the number of COVID-19 tweets, but the sentiment of the COVID-19 tweets did not have an obvious pattern in the United States compared to the United Kingdom. This is probably a consequence of emotional numbness when the United States had much more COVID-19 cases than the United Kingdom.

## Limitations

There were several limitations in our study. First, although Twitter is one of the most popular social media platforms [27], the Twitter users may not represent the whole population. Second, our study only focused on Twitter data from March 6 to April 2, 2020. COVID-19 is an ongoing pandemic and public reactions towards COVID-19 might evolve after April 2, 2020. Third, the geographic information used in our study might have some biases as the user geolocation in their profile could be inaccurate.

## Conclusion

By analyzing COVID-19 related tweets from March 6 to April 2, 2020 in the United Kingdom and the United States, we showed the differences in the public attitudes towards COVID-19 in different countries in a timely manner, which might correlate with the number of COVID-19 cases and some important policies/news related to COVID-19. Our study provides some evidence about the correlation between the severity of COVID-19 pandemic and the public attitudes towards COVID-19, especially how different policies from different countries affect the public attitudes towards the COVID-19 pandemic.

## Data Availability

The number of confirmed COVID-19 cases in the United Kingdom and the United States from March 6, 2020 to April 2, 2020 were obtained from the 1Point3Acres website. The Twitter dataset used for this study was generated by a crawler using the Twitter streaming API from March 6, 2020 to April 2, 2020.

## Acknowledgments

The project described in this publication was supported by the University of Rochester Clinical and Translational Science Award UL1 TR002001 from the National Center for Advancing Translational Sciences of the National Institutes of Health (DL).

## Reference

[1] “WHO | Novel Coronavirus – China,” WHO. http://www.who.int/csr/don/12-january-2020-novel-coronavirus-china/en/ (accessed Jun. 19, 2020).

[2] “Coronavirus.” https://www.who.int/westernpacific/health-topics/coronavirus (accessed Jun. 16, 2020).

[3] “What is pandemic? Why did WHO just declare one? - The Washington Post.” https://www.washingtonpost.com/health/2020/03/11/who-declares-pandemic-coronavirus-disease-covid-19/ (accessed Jun. 19, 2020).

[4] “Global COVID-19 Tracker & Interactive Charts | Real Time Updates & Digestable Information for Everyone | 1Point3Acres.” https://coronavirus.1point3acres.com (accessed Jun. 16, 2020).

[5] J. H. Jones and M. Salathé, “Early Assessment of Anxiety and Behavioral Response to Novel Swine-Origin Influenza A(H1N1),” PLOS ONE, vol. 4, no. 12, p. e8032, Dec. 2009, doi:10.1371/journal.pone.0008032.

[6] “Q1-2019-Slide-Presentation.pdf.” Accessed: Jun. 17, 2020. [Online]. Available: https://s22.q4cdn.com/826641620/files/doc_financials/2019/q1/Q1-2019-Slide-Presentation.pdf.

[7] H. W. Park, S. Park, and M. Chong, “Conversations and Medical News Frames on Twitter: Infodemiological Study on COVID-19 in South Korea,” J. Med. Internet Res., vol. 22, no. 5, p. e18897, 2020, doi:10.2196/18897.

[8] R. Kouzy et al., “Coronavirus Goes Viral: Quantifying the COVID-19 Misinformation Epidemic on Twitter,” Cureus, vol. 12, no. 3, doi:10.7759/cureus.7255.

[9] W. Ahmed, J. Vidal-Alaball, J. Downing, and F. L. Seguí, “COVID-19 and the 5G Conspiracy Theory: Social Network Analysis of Twitter Data,” J. Med. Internet Res., vol. 22, no. 5, p. e19458, 2020, doi:10.2196/19458.

[10] N. Aguilar-Gallegos, L. E. Romero-García, E. G. Martínez-González, E. I. García-Sánchez, and J. Aguilar-Ávila, “Dataset on dynamics of Coronavirus on Twitter,” Data Brief, vol. 30, p. 105684, Jun. 2020, doi:10.1016/j.dib.2020.105684.

[11] J. Gao et al., “Mental health problems and social media exposure during COVID-19 outbreak,” PLOS ONE, vol. 15, no. 4, p. e0231924, Apr. 2020, doi:10.1371/journal.pone.0231924.

[12] Rufai S. R. and Bunce C., “World leaders’ usage of Twitter in response to the COVID-19 pandemic: a content analysis,” J. Public Health, doi:10.1093/pubmed/fdaa049.

[13] “Overview.” https://developer.twitter.com/en/docs/tweets/filter-realtime/overview (accessed Jun. 19, 2020).

[14] C. J. Hutto, cjhutto/vaderSentiment. 2020.

[15] “gensim: topic modelling for humans.” https://radimrehurek.com/gensim/models/ldamulticore.html (accessed Jun. 19, 2020).

[16] “Natural Language Toolkit — NLTK 3.5 documentation.” https://www.nltk.org/ (accessed Jun. 19, 2020).

[17] “spaCy · Industrial-strength Natural Language Processing in Python.” https://spacy.io/ (accessed Jun. 19, 2020).

[18] “pyLDAvis — pyLDAvis 2.1.2 documentation.” https://pyldavis.readthedocs.io/en/latest/readme.html (accessed Jun. 19, 2020).

[19] R. Partington and P. Walker, “Rishi Sunak promises to guarantee £330bn loans to business,” The Guardian, Mar. 17, 2020.

[20] “A Guide to State Coronavirus Reopenings and Lockdowns - WSJ.” https://www.wsj.com/articles/a-state-by-state-guide-to-coronavirus-lockdowns-11584749351?mod=theme_coronavirus-ribbon (accessed Jun. 19, 2020).

[21] “Boris Johnson announces ‘extraordinary’ closure of UK’s pubs and restaurants in coronavirus shutdown.” https://www.telegraph.co.uk/politics/2020/03/20/boris-johnson-announces-extraordinary-closure-uks-pubs-restaurants/ (accessed Jun. 19, 2020).

[22] “Coronavirus: Prince Charles tests positive but ‘remains in good health’ - BBC News.” https://www.bbc.com/news/uk-52033845 (accessed Jun. 19, 2020).

[23] “Coronavirus: Prime Minister Boris Johnson tests positive - BBC News.” https://www.bbc.com/news/uk-52060791 (accessed Jun. 19, 2020).

[24] T. Nguyen, D. Phung, B. Adams, and S. Venkatesh, “Event extraction using behaviors of sentiment signals and burst structure in social media,” Knowl. Inf. Syst. Lond., vol. 37, no. 2, pp. 279–304, Nov. 2013, doi:http://dx.doi.org.ezp.lib.rochester.edu/10.1007/s10115-012-0494-9.

[25] F. Dzogang, S. Lightman, and N. Cristianini, “Diurnal variations of psychometric indicators in Twitter content.,” PLoS ONE, vol. 13, no. 6, p. e0197002, Jan. 2018, doi:10.1371/journal.pone.0197002.

[26] S. A. Golder and M. W. Macy, “Diurnal and Seasonal Mood Vary with Work, Sleep, and Daylength Across Diverse Cultures,” Science, vol. 333, no. 6051, pp. 1878–1881, Sep. 2011, doi:10.1126/science.1202775.

[27] “Global social media ranking 2019,” Statista. https://www.statista.com/statistics/272014/global-social-networks-ranked-by-number-of-users/ (accessed Jun. 19, 2020).

